# Lateral hypothalamus drives early-onset sleep alterations in amyotrophic lateral sclerosis

**DOI:** 10.1101/2024.08.21.24312343

**Authors:** Simon J. Guillot, Christina Lang, Marie Simonot, Antje Knehr, Geoffrey Stuart-Lopez, Patrick Weydt, Johannes Dorst, Katharina Kandler, Jan Kassubek, Laura Wassermann, Caroline Rouaux, Sebastien Arthaud, Pierre-Herve Luppi, Francesco Roselli, Albert C. Ludolph, Luc Dupuis, Matei Bolborea

## Abstract

Lateral hypothalamic neurons producing melanin-concentrating hormone (MCH) and orexin/hypocretin are involved in sleep regulation. Both MCH and orexin neurons are altered in amyotrophic lateral sclerosis (ALS), the most common adult-onset motor neuron disease. However, sleep alterations are currently poorly characterized in ALS, and could represent either early symptoms or late consequences of disease progression. Here, we characterized sleep architecture using polysomnography in cohorts of both early ALS patients without respiratory impairment and presymptomatic carriers of mutations leading to familial ALS. We observed prominent sleep alterations, including increased wake and decreased deep sleep (non-rapid eye movement—NREM3) in both cohorts, which were replicated in two mouse models of familial ALS, *Sod1^G86R^* and *Fus^ΔNLS/+^* mice. Importantly, altered sleep structure in mice was fully rescued by *per os* administration of a dual-orexin receptor antagonist, and partially rescued by intracerebroventricular MCH supplementation. Thus, our study shows the existence of a primary sleep alteration in ALS, driven by abnormal MCH and orexin signalling.

**One Sentence Summary:** Amyotrophic lateral sclerosis is a tragic uncurable motor neuron disease, in this study we decribed for the first time sleep alterations in symptomatic patients and healthy gene carrier which can be reverted by *per os* administartion of a dual-orexin receptor antagonist in preclinical models.

## INTRODUCTION

Amyotrophic lateral sclerosis (ALS), the most common adult-onset motor neuron disorder, is a fatal disease, mostly leading to death through progressive paralysis and respiratory insufficiency within two to three years after the onset of symptoms (*1*). Depending on populations, the median age of onset for sporadic ALS is 55 to 65 years of age (*2*). Sporadic ALS (90-95%) accounts for the majority of cases, while the remaining 5-10% are hereditary and referred to as familial ALS (*3*). More than 30 different genes have been associated with familial ALS, with mutations in *C9ORF72, SOD1*, *TARDBP* and *FUS* being the most frequent causes of familial ALS (*4–8*).

While ALS is clinically defined as the simultaneous degeneration of lower motor neurons - in the brainstem and spinal cord - and of upper motor neurons - in the motor cortex -, recent years have demonstrated that ALS broadly affects multiple brain functions and that non-motor brain regions are also affected, including the hypothalamus. Hypothalamic atrophy was observed in ALS patients and presymptomatic gene carriers using magnetic resonance imaging (MRI) (*9*), a finding confirmed by several other groups (*10–12*). Furthermore, there were functional abnormalities in the response of hypothalamus-controlled functions to drugs or fasting, both in ALS patients and mouse models (*13*). Recently, we observed prominent neurodegeneration and ALS-related pathology (*i*.*e*., TDP-43 aggregates) in the lateral hypothalamic area (LHA) (*14*), consistent with recent study (*15*). In the LHA the key neuronal populations, melanin-concentrating hormone (MCH) neurons and orexin/hypocretin (ORX) neurons, are critical in sleep regulation (*16–18*), and both neuronal populations appear affected in ALS patients (*14, 19, 20*).

Degeneration of MCH and ORX neurons in ALS raises the possibility that sleep could be altered in ALS. Only few studies have investigated sleep in ALS. Overall, in studies available to date more than half of ALS patients report poor subjective sleep quality and a few polysomnography studies suggest altered sleep architecture among ALS patients (*21–23*). However, previous studies included advanced ALS patients that develop respiratory insufficiency due to disease progression and are therefore not suitable for determining whether sleep alterations occur independently, and possibly before, motor symptoms. Thus, it remains unknown whether sleep is primarily affected in ALS (*24*).

In the current study, we characterized sleep structure in a cohort of 56 early ALS patients versus 41 healthy individuals and a second cohort including 35 presymptomatic gene carriers of ALS mutations (part of 62 healthy first-degree relatives of confirmed fALS patients, genetically screened for susceptibility genes). We controlled for respiratory involvement during sleep by transcutaneous capnometry. We observed early prominent sleep alterations in both cohorts and replicated a major part of these observations in two mouse models, S*OD1^G86R^* and *Fus^ΔNLS/+^*. Furthermore, inhibiting ORX signalling or intracerebroventricular supplementation with MCH is sufficient to rescue sleep alterations in both ALS mouse models.

Collectively, our results demonstrate that sleep alterations are early and severe in ALS and are causally related to abnormalities in MCH and ORX balance signalling.

## RESULTS

### Early ALS patients without respiratory impairment show altered sleep macroarchitecture

To investigate the prevalence of sleep alterations in early symptomatic ALS patients, we designed a prospective cohort. Three main exclusion criteria were the presence of (i) sleep apnoea, (ii) periodic limb movements in sleep and/or (iii) nocturnal hypercapnia (see **Figure 1a**). In all, 56 patients and 41 age- and sex-matched controls were screened, and 33 ALS patients and 32 controls met the inclusion criteria (**Figure 1a**, **Table 1**).

The group of ALS patients versus controls showed no significant differences in age, sex distribution, and body mass index. The cohort is described in **Table 1**. In the ALS patient group, function was recorded with the ALSFRS-R (Amyotrophic Lateral Sclerosis Functional Rating Scale Revised) at the time of polysomnography (*25*). The resulting mean of 40.51 (±0.78) indicates that this is a group of patients in early stages of the disease.

Analysis of questionnaires did not reveal any difference in subjective sleep between ALS patients and controls (**Figure S1**). Polysomnography results were analysed using YASA deep learning algorithm (*26*), in addition to the manual evaluation of the recordings based on the evaluation criteria of the American Academy of Sleep Medicine, Version 2.6 (*27*), each over a time-window of 6 hours after lights off. Hypnograms obtained using deep learning sleep analysis were highly concordant (91.6% ±2.58; n=8, **Figure S2**) to manually determined hypnograms (**Figure 1b**). ALS patients showed an increased total sleep time and increased sleep latency (**Figure 1c-d**). Sleep macroarchitecture in ALS patients was severely altered (**Table S2**) with increased percentage of wake (**Figure 1e**), of rapid eye movement (REM) sleep (**Figure 1f**) and decreased non rapid eye movement (NREM) sleep (**Figure 1g**). Decreased NREM sleep was mostly due to a strong decrease in NREM3 (deep sleep) and NREM2, while NREM1 (light sleep) was preserved (**Figure 1h-j**). Principal component analysis (PCA) using sleep parameters and age showed a complete segregation of ALS patients from controls (**Figure 1k**). Thus, sleep macroarchitecture is affected in ALS patients, in the absence of confounding respiratory insufficiency.

**Table 1.**
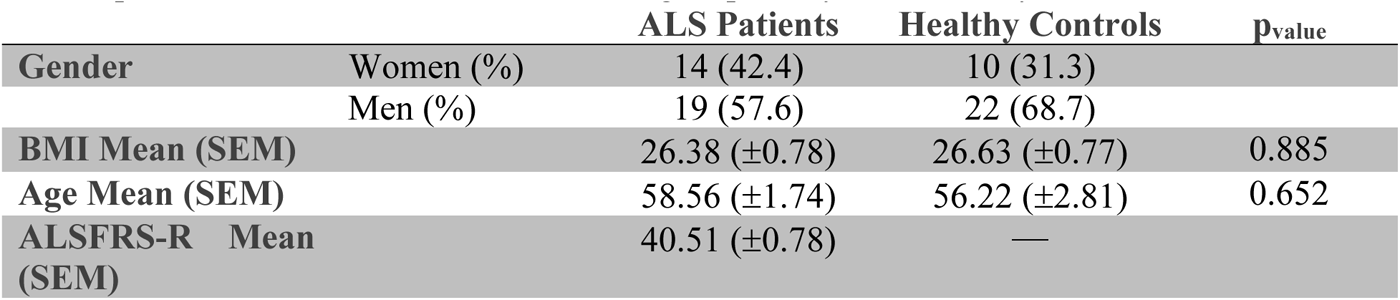
Descriptive statistics of the study population of ALS patients and healthy controls. SEM: standard error of means; BMI: body mass index; ALSFRS-R: Amyotrophic Lateral Sclerosis Functional Rating Scale Revised; n.s. p_value_>0.05, non-parametric Kruskal-Wallis’ test.

**Figure 1.**
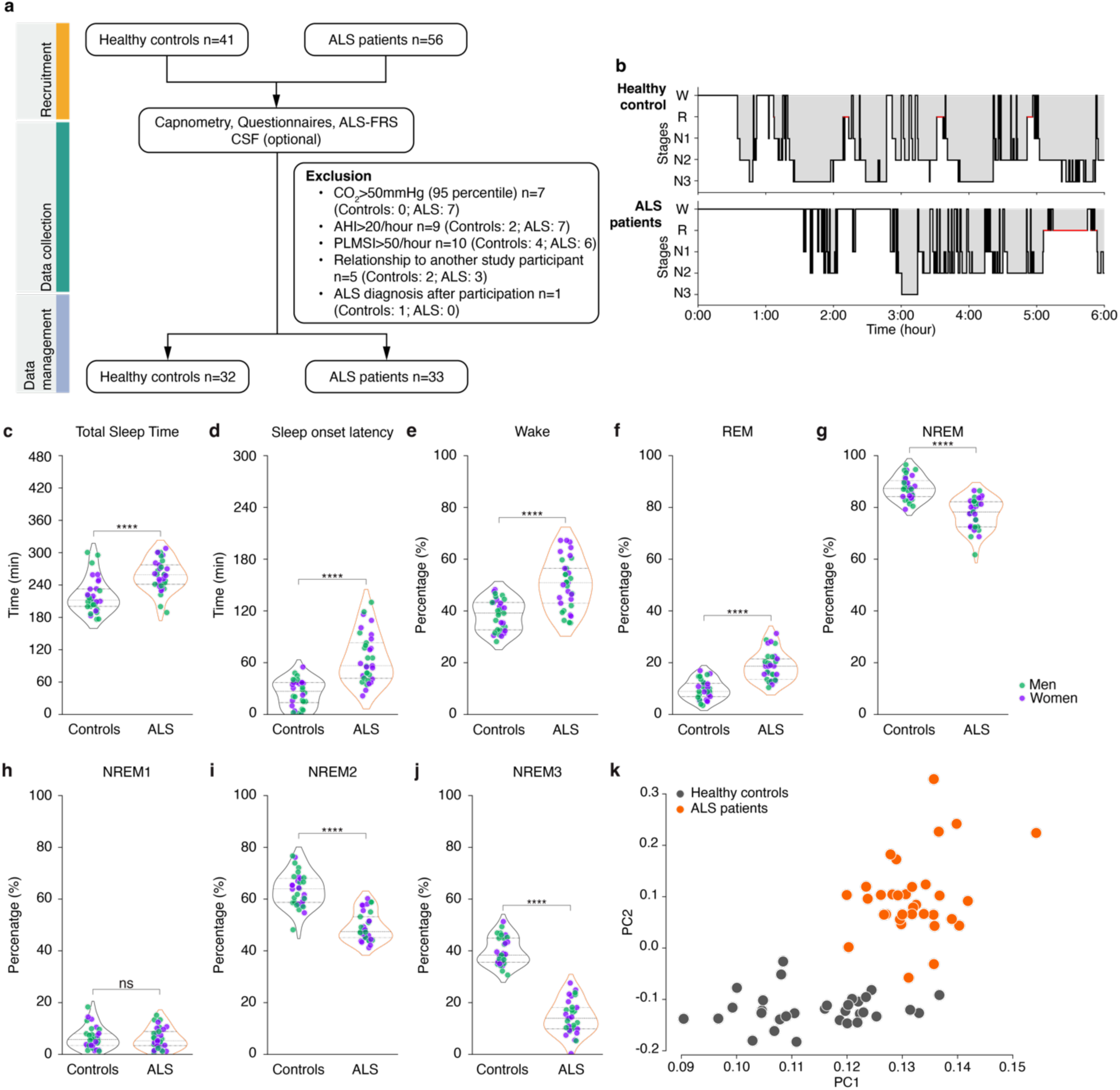
Sleep alterations in early ALS patients. (**a**) Flow chart of the study. (**b**) Representative hypnograms of a healthy control and one sporadic ALS patient over a 6h period (1 epoch=30 seconds). (**c**) Total sleep time (total duration of REM, NREM1, NREM2 and NREM3 in the sleep period time). (**d**) Sleep onset latency (latency to the first epoch of any sleep). (**e**-**j**) Percentage of wake (**e**), REM (**f**), NREM (**g**), NREM1 (**h**), NREM2 (**i**) and NREM3 (**j**). (**k**) PCA analysis of ALS patients versus healthy controls using sleep parameters and age. In all panels, men are shown in green and women in purple. Corrected p_value_ are shown. **** adj. p_value_<0.0001, independent Student’s t-test with Welch’s t-test correction; sex effect adj. p_value_=0.4296. Data are presented as median and interquartile ranges.

### Presymptomatic ALS gene carriers show similarly altered sleep macroarchitecture

While our first cohort analysis indicated the presence of sleep alterations in early-stage ALS patients even in the absence of hypercapnia, it could not ascertain that sleep alterations are already present prior to motor symptoms. Therefore, we drew on a second prospective cohort study that consisted of presymptomatic ALS gene carriers, using the same inclusion and exclusion criteria (see **Figure 2a**). A total of 62 first-degree relatives of fALS patients were initially screened, of whom 35 individuals had a positive fALS gene test result. Participants were blinded to their test results but could opt to undergo counselling if they wanted to learn their mutation status in accordance with German legislation. Of these, 28 individuals met the above inclusion criteria (*SOD1* n=7; *C9ORF72* n=12). The first-degree relatives with a negative genetic test constituted our control group (fALS controls; 27 screened, 19 meeting inclusion criteria) (**Figure 2a**, **Table 2**). Like early ALS patients, sleep questionnaires did not reveal alterations in subjective sleep in ALS gene carriers (**Figure S3**). Nevertheless, presymptomatic ALS gene carriers, like ALS patients, already exhibit macroarchitectural alterations of their sleep pattern. Partly varying results were observed depending on the mutation. Total sleep time was decreased in *SOD1* gene carriers but unchanged in *C9ORF72* gene carriers (**Figure 2b-c**), while only *C9ORF72* gene carriers displayed longer sleep onset latency (**Figure 2d**). Interestingly, and like in early symptomatic ALS patients, we observed an increased percentage of wake phases (**Figure 2e**) and of REM sleep (**Figure 2f**), and decreased NREM sleep (**Figure 2g**) in *C9ORF72* gene carriers, which was mostly due to decreased NREM2 and -3 (**Figure 2h-j**). *SOD1* gene carriers displayed intermediate sleep changes with increased wake and decreased NREM2/3 but normal REM sleep. Consistent with this, PCA revealed a more defined segregation of *C9ORF72* gene carriers from controls than *SOD1* gene carriers (**Figure 2k**). These results show that sleep alterations are present in individuals carrying ALS risk genes, in particular *C9ORF72,* many years before expected motor symptom onset.

**Table 2.** Descriptive statistics of the study population of fALS participants. SEM: standard error of means; BMI: body mass index; ALSFRS-R: Amyotrophic Lateral Sclerosis Functional Rating Scale Revised; n.s. p_value_>0.05, non-parametric Kruskal-Wallis’ test.

| | | fALS Gene Carriers | fALS Controls | $p_{\text{value}}$ |
| --- | --- | --- | --- | --- |
| Gender | Women (%) | 20 (71.4) | 14 (73.7) |  |
|  | Men (%) | 8 (28.6) | 5 (26.3) |  |
| BMI Mean (SEM) | | 26.88 ($\pm 0.89$) | 26.06 ($\pm 1.59$) | 0.72 |
| Age (SEM) | | 40.66 ($\pm 2.88$) | 41.84 ($\pm 2.67$) | 0.81 |

**Figure 2.**
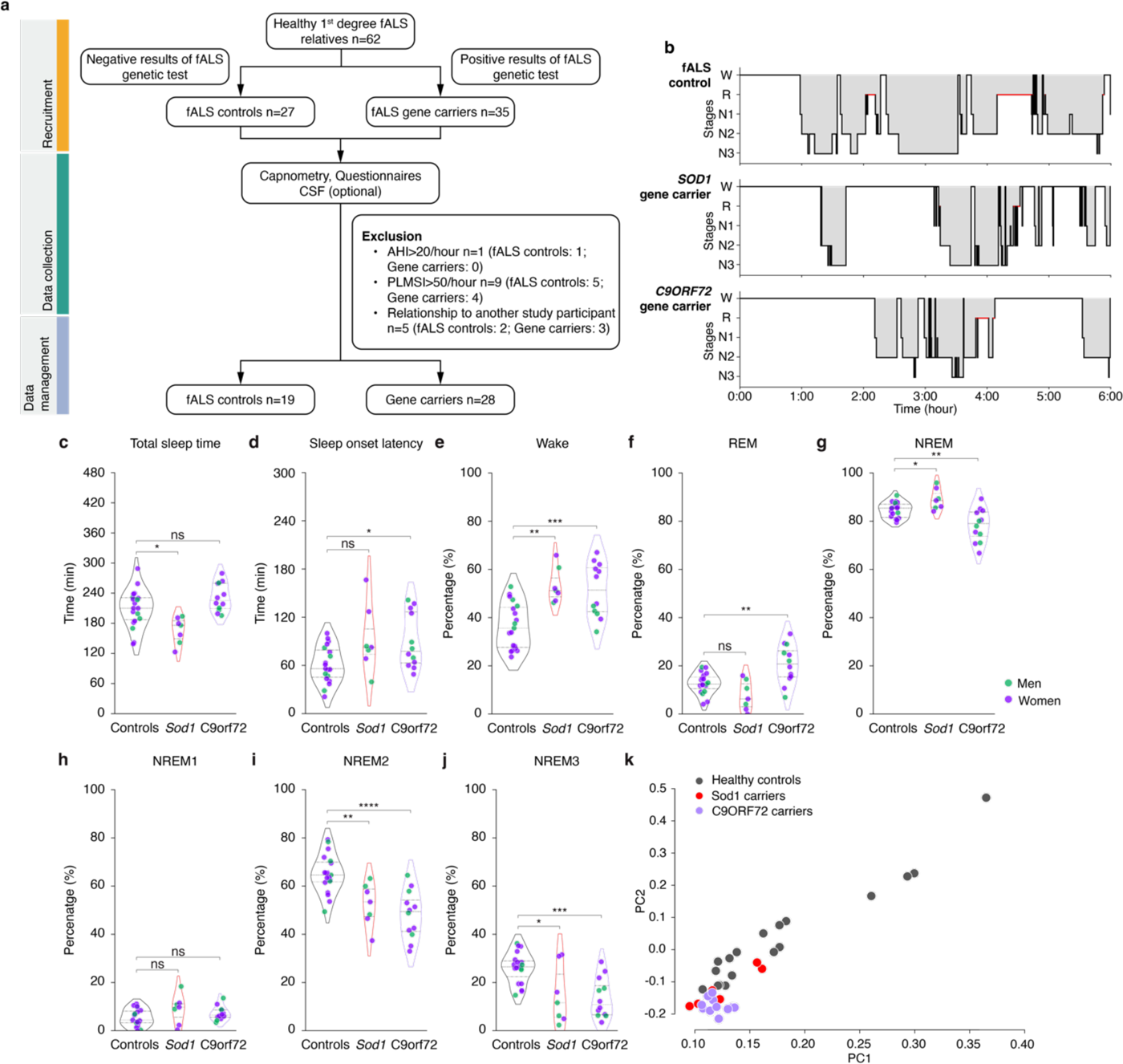
Sleep alterations in presymptomatic ALS gene carriers. (**a**) Flow chart of the study. (**b**) Representative hypnograms of a fALS control, one *SOD1* ALS gene carrier and one *C9ORF72* gene carrier over a 6h period (1 epoch=30 seconds). (**c**) Total sleep time (total duration of REM, NREM1, NREM2 and NREM3 in the sleep period time). (**d**) Sleep onset latency (latency to the first epoch of any sleep). (**e**-**j**) Percentage of wake (**e**), REM (**f**), NREM (**g**), NREM1 (**h**), NREM2 (**i**) and NREM3 (**j**). (**k**) PCA analysis of presymptomatic *SOD1, C9ORF72* gene carriers and fALS controls using sleep parameters. In all panels, men are shown in green and women in purple. * adj. p_value_<0.05, One-way ANOVA with one-step Bonferroni correction; sex effect adj. p_value_=0.4096. Data are presented as median and interquartile ranges. Corrected p_value_ are shown.

### Two mouse models of ALS show altered sleep macroarchitecture

The presence of altered sleep macroarchitecture in ALS patients and presymptomatic gene carriers prompted us to investigate sleep patterns in transgenic ALS mouse models. We used two models with different ALS-causing genes and vastly divergent disease progression. Transgenic expression of G86R mutation in SOD1 leads to severe, fast progressing motor symptoms (*28, 29*), while knock-in expression of a C-terminally truncated FUS protein leads to mild, late-onset motor neuron disease (*30, 31*). To characterize sleep patterns, we implanted intra-cortical electrodes at 60 days of age and performed electrocorticography 5-6 days after surgery (**Figure 3a**). Quantification of sleep states through manual analysis, or automated sleep analysis using NeuroScore, showed an overall concordance of 94.49% **±**2.27 (n=6, **Figure S4**) and hypnograms obtained were highly similar in a pilot experiment. We thus relied on automated sleep analysis for further experiments. Hypnograms of *Sod1^G86R^* mice showed increased wake, decreased NREM and decreased REM sleep (**Figure 3b-e**) at 75 days of age, *i.e.,* before the appearance of motor defects and weight loss at 90 days of age. In contrast, there were no alterations in sleep patterns at 3 months of age in *Fus^ΔNLS/+^* mice (**Figure 3f-i**), but 10 months old *Fus^ΔNLS/+^* mice also showed increased wake, decreased NREM and decreased REM sleep (**Figure 3j-m**). In both mouse models, the sleep phenotype was consistently observed in both male and female mice. Thus, ALS mouse models recapitulate the early sleep alterations observed in humans, except for percentage of REM sleep, which is increased in humans and decreased in mice.

**Figure 3.**
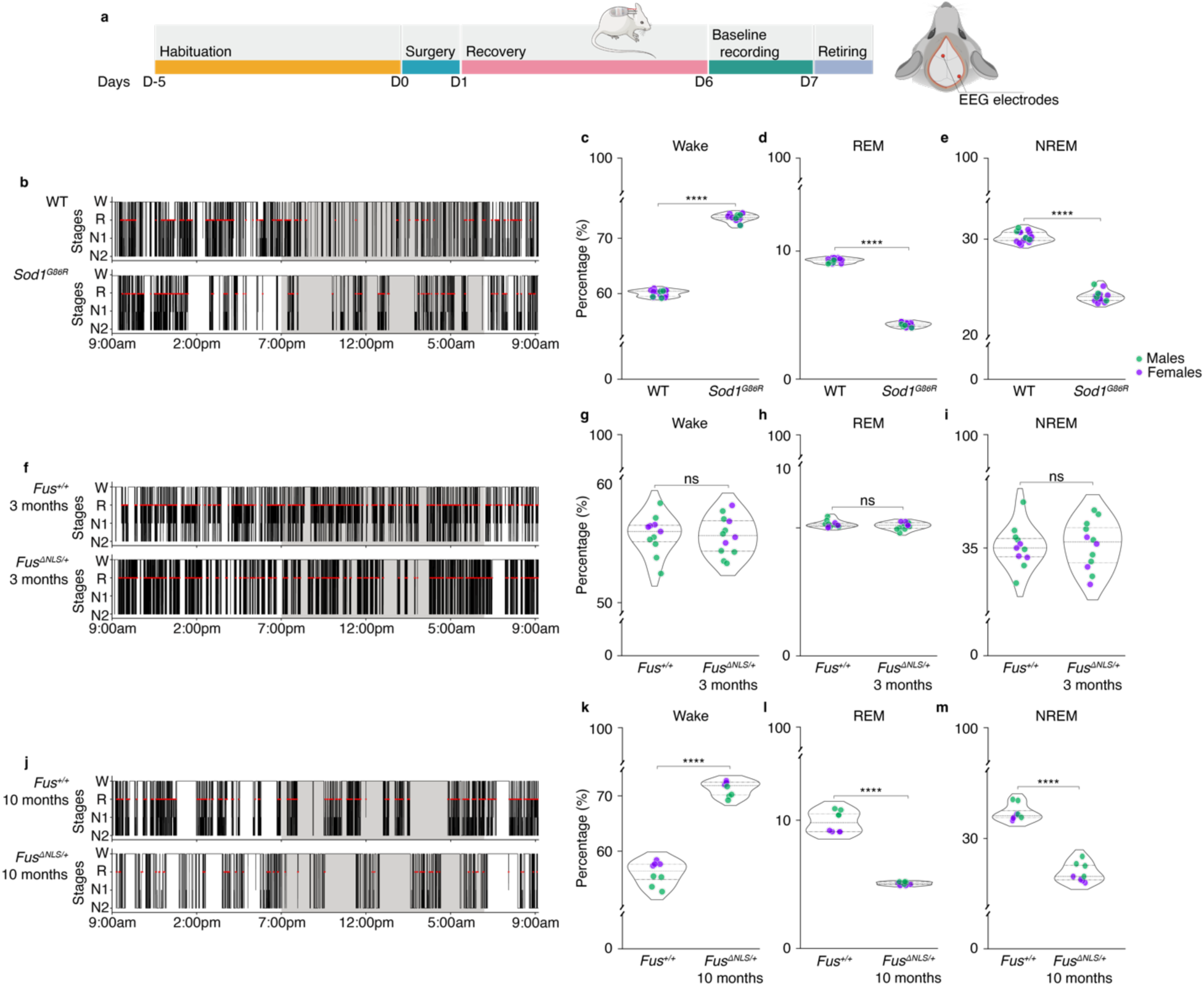
Sleep alterations in *Sod1^G86R^* and *Fus^ΔNLS/+^* mice. (**a**) Experimental design. (**b**-**e**) Representative hypnograms of *Sod1^G86R^* mice and their wild-type non transgenic (WT) littermates at 75 days of age (prior to motor symptom onset) (**b**), and quantification of wake (**c**), REM (**d**) and NREM (**e**) *Sod1^G86R^*mice: n=14; 10 females and 4 males; WT: n=18; 14 females and 4 males. (**f**-**i**) Representative hypnograms of *Fus*^ΔNLS/+^ mice and their WT littermates (*Fus*^+/+^) at 3 months of age (**f**), and quantification of wake (**g**), REM (**h**) and NREM (**i**) *Fus*^ΔNLS/+^ mice: n=12; 4 females and 8 males; *Fus*^+/+^: n=11; 4 females and 7 males. (**j**-**m**) Representative hypnograms of *Fus*^ΔNLS/+^ mice and their WT littermates (*Fus*^+/+^) at 10 months of age (**j**), and quantification of wake (**g**), REM (**h**) and NREM (**i**) *Fus*^ΔNLS/+^ mice: n=8; 4 females and 4 males; *Fus*^+/+^: n=8; 4 females and 4 males. Recordings were performed over a 24h period (1 epoch=5 seconds). Grayed areas correspond to the active phase, when lights were off, whereas white areas indicate the non-active phase, where lights are on. **** adj. p_value_<0.0001, independent Student’s t-test with Welch’s t-test correction; sex effect *Sod1*^G86R^ adj. p_value_=0.6302, *Fus*^ΔNLS/+^ 3 months adj. p_value_=0.6708, *Fus*^ΔNLS/+^ 10 months adj. p_value_=0.0953. Data are presented as median and interquartile ranges. Corrected p_value_ are shown.

### Sleep alterations in mouse models are partially rescued by MCH

Since MCH neurons degenerate in ALS and MCH promotes sleep - in particular REM sleep (*17, 32, 33*) - we hypothesized that MCH loss could contribute to the observed sleep alterations. To test this, we implanted an intracerebroventricular (i.c.v.) osmotic pump in parallel with intra-cortical electrodes for electrocorticography (EEG) (**Figure 4a**). Pumps were filled with either the vehicle or MCH (*14*). While vehicle-treated *Sod1^G86R^* mice showed similar sleep alterations as untreated *Sod1^G86R^* mice, MCH treatment fully rescued decreased REM sleep in *Sod1^G86R^* mice, and partially rescued increased wake (**Figure 4b-e**). However, MCH infusion exacerbated the NREM sleep deficit (**Figure 4b-e**). Similarly, partial normalizing effects of MCH on sleep architecture were also observed in *Fus^ΔNLS/+^* mice (**Figure 4f-i**). MCH supplementation could rescue REM sleep alterations as well as NREM and wake alterations in ALS mouse models.

**Figure 4.**
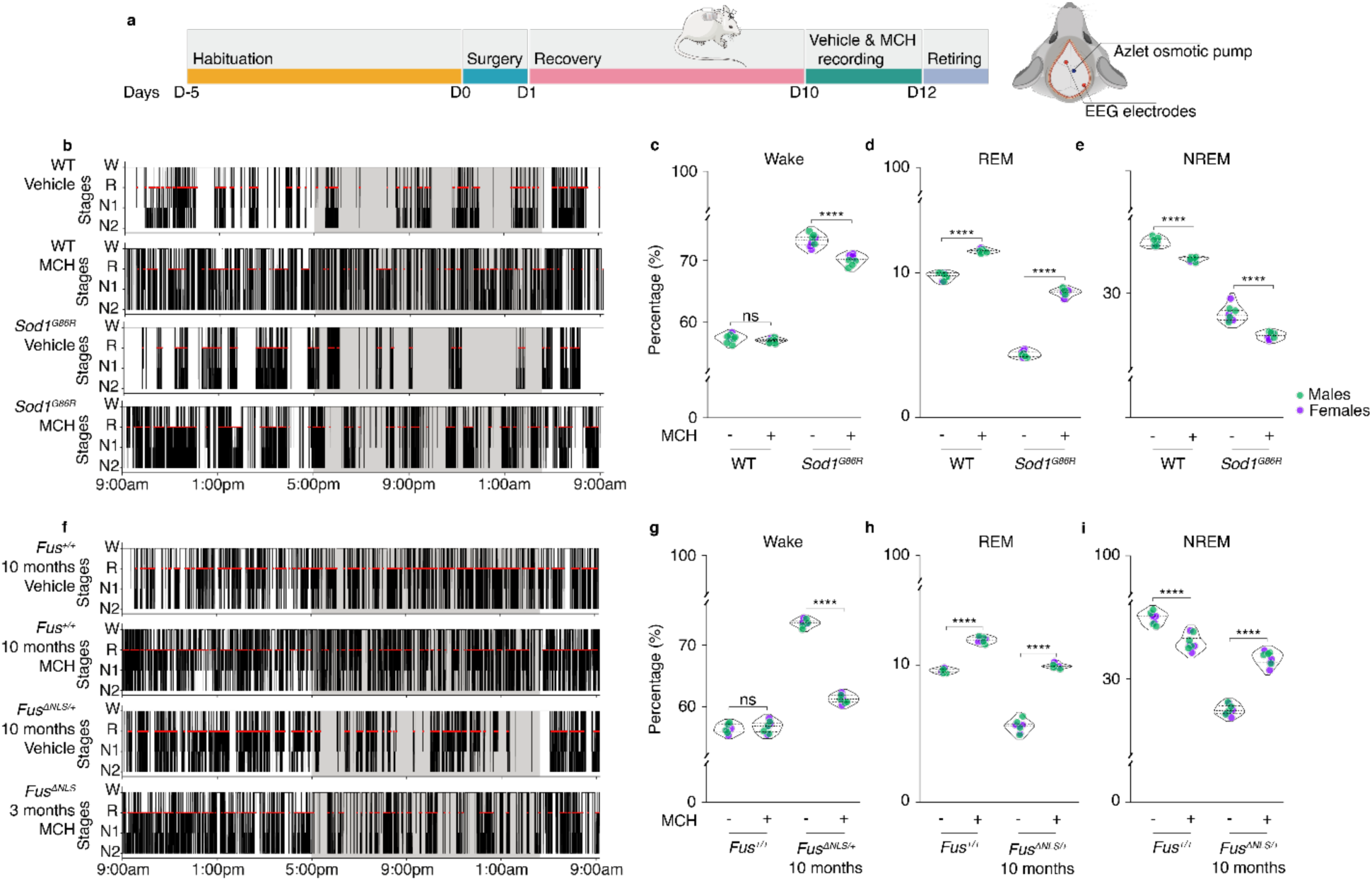
MCH partially rescued sleep alterations in *Sod1^G86R^* and *Fus^ΔNLS/+^* mice. (**a**) Experimental design. (**b**-**e**) Representative hypnograms of *Sod1^G86R^* mice and their WT non transgenic littermates at 75 days of age (prior to motor symptom onset) (**b**), and quantification of wake (**c**), REM (**d**) and NREM (**e**) *Sod1^G86R^* mice: n=8; 4 females and 4 males; WT: n=8; 2 females and 6 males. (**f**-**i**) Representative hypnograms of *Fus^ΔNLS/+^* mice and their WT littermates (*Fus^+/+^*) at 10 months of age (**f**), and quantification of wake (**g**), REM (**h**) and NREM (**i**) *Fus^ΔNLS/+^*mice: n=8; 4 females and 4 males; *Fus^+/+^*: n=8; 4 females and 4 males. Recordings were performed over a 24h period (1 epoch=5 seconds). Grayed areas correspond to the active phase, when lights were off, whereas white areas indicate the non-active phase, where lights are on. *** adj. p_value_<0.001, Two-way ANOVA with one-step Bonferroni correction; genotype effect *Sod1*^G86R^ adj. p_value_<0.0001, *Fus*^ΔNLS/+^ 10 months adj. p_value_<0.0001; sex effect *Sod1*^G86R^ adj. p_value_=0.9187, *Fus*^ΔNLS/+^ 10 months adj. p_valu_e=0.1442. Data are presented as median and interquartile ranges. Corrected p_value_ are shown.

### Sleep alterations in mouse models are fully rescued by a dual orexin receptor antagonist

ORX and MCH neurons play partially antagonistic roles in sleep/wake regulation. We thus hypothesized that ORX neurons could also be involved in the observed sleep alterations, *e.g.*, through overactive orexinergic tone. We first characterized ORX neuronal counts in presymptomatic *Sod1^G86R^*(75 days of age) and *Fus^ΔNLS/+^* (10 months of age) mice but did not observe loss of this neuronal population in these mouse models (**Figure S5**).

Dual orexin receptor antagonists (DORA), such as the FDA-approved drug Suvorexant, can acutely inhibit orexin signalling (*34, 35*). We thus administered either Suvorexant or its vehicle in mouse models equipped with EEG cortical electrodes (**Figure 5a**). As mice are nocturnal animals, they are at rest/sleep during the day (“light phase”) and active during the night (“dark phase”). Acute administration of Suvorexant at onset of light phase, was able to rescue sleep alterations in *Sod1^G86R^* mice. In particular, it normalized wake, REM and NREM sleep to levels similar to wild-type untreated animals (**Figure 5b-e**), similar results were obtained in 10 months old *Fus^ΔNLS/+^*mice (**Figure 5f-i**). This effect was observed in both male and female mice. Importantly, and contrary to MCH, Suvorexant was able to fully rescue all sleep alterations, suggesting a broader involvement of ORX than of MCH in sleep alterations in ALS versus vehicle administration.

**Figure 5.**
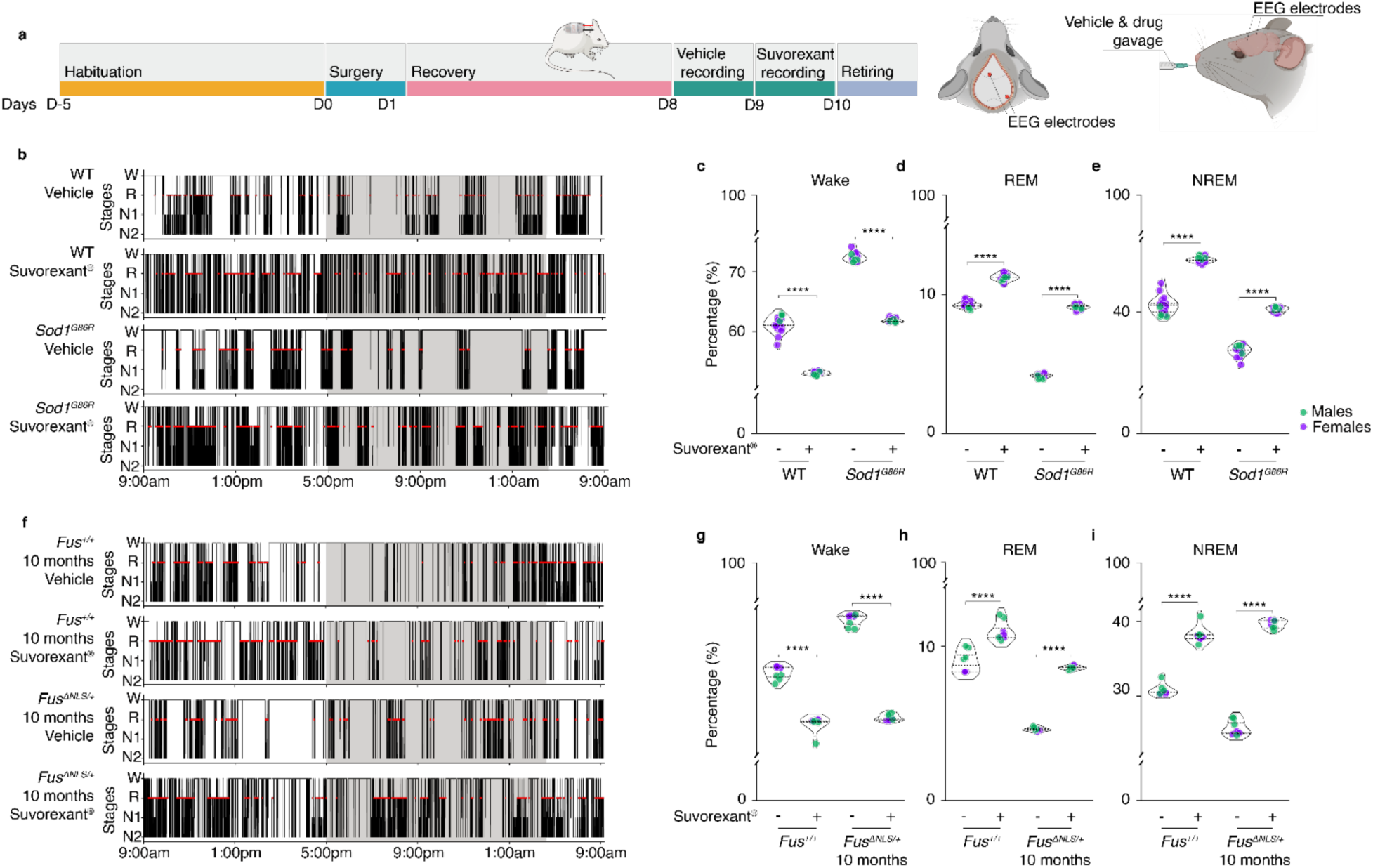
Rescued sleep alterations by repurposing Suvorexant in *Sod1^G86R^* and *Fus^ΔNLS/+^* mice. (**a**) Experimental design. (**b**-**e**) Representative hypnograms of *Sod1^G86R^* mice and their WT non transgenic (WT) littermates after administration of the vehicle solution or Suvorexant at 75 days of age (prior to motor symptom onset), (**b**), and quantification of wake (**c**), REM (**d**) and NREM (**e**) *Sod1^G86R^*mice: n=14; 10 females and 4 males; WT: n=18; 14 females and 4 males. (**f**-**i**) Representative hypnograms of *Fus^ΔNLS/+^* mice and their WT littermates (*Fus^+/+^*) at 10 months of age after administration of the vehicle solution or Suvorexant (**f**), and quantification of wake (**g**), REM (**h**) and NREM (**i**) *Fus^ΔNLS/+^* mice: n=8; 4 females and 4 males; *Fus^+/+^*: n=8; 4 females and 4 males. Recordings were performed over a 24h period (1 epoch=5 seconds). Grayed areas correspond to the active phase, when lights were off, whereas white areas indicate the non-active phase, where lights are on. **** adj. p_value_<0.0001, Two-way ANOVA with one-step Bonferroni correction; genotype effect *Sod1*^G86R^ adj. p_value_<0.0001, *Fus^ΔNLS/+^* 3 months adj. p_value_=0.0116, *Fus^ΔNLS/+^* 10 months adj. p_value_<0.0001; sex effect *Sod1*^G86R^ adj. p_value_=0.4722, *Fus^ΔNLS/+^* 3 months adj. p_value_=0.7640, *Fus^ΔNLS/+^* 10 months adj. p_value_=0.2218. Data are presented as median and interquartile ranges. Corrected p_value_ are shown.

## DISCUSSION

Alterations in sleep are a hallmark of multiple neurodegenerative disorders as well as of normal aging (*36*), and ALS is not an exception. Several previous studies have shown the occurrence of sleep alterations in ALS patients (*23, 37*).

However, available studies to date included patients with manifest disease, thus with possibly ongoing progressive respiratory impairment as an effect of disease progression (*38*). Consistently, sleep alterations were usually correlated to disease progression in these studies. Thus, these studies could not determine whether sleep alterations were secondary to disease progression or pre-existed the motor defects. Our current study provides complementary evidence, both in patients and in mouse models, that sleep alterations are an early phenomenon in ALS which precedes respiratory impairment and even motor symptoms.

To ascertain that sleep alterations are independent of respiratory decline, we performed two clinical cohort studies in different populations. First, we prospectively included early ALS patients and pre-specified in our exclusion criteria multiple outcomes related to sleep disorders, possibly altering sleep architecture, including nocturnal hypercapnia. This ensured that all included participants had normal respiratory function. We uncovered striking alterations in sleep macroarchitecture of ALS patients, in particular increased sleep onset latency, decreased deep sleep (NREM2 and NREM3), and increased wake after sleep onset, none of which were self-reported by the participants. While this first study showed the early occurrence of sleep alterations in patients with manifest disease, it was impossible to exclude that neurological deficits including ongoing motor symptoms might have influenced sleep alterations.

To circumvent this limitation, we performed an analogous prospective cohort study in presymptomatic ALS gene carriers. Indeed, we observed a similar, yet milder, sleep phenotype in this population of presymptomatic gene carriers. *C9ORF72* gene carriers showed alterations in the same direction as ALS patients for sleep onset latency, deep sleep (NREM2 and NREM3), and wake after sleep onset. *SOD1* gene carriers only showed decreased deep sleep and increased wake after sleep onset, consistent with a milder clinical picture in *SOD1* as compared to *C9ORF72* ALS patients.

Consistent with the two cohort studies, we observed almost identical sleep alterations in two transgenic mouse models of ALS. In mice expressing mutant SOD1, that are characterized by rapid disease progression and appearance of symptoms at about 3 months of age, we observed increased wake and decreased NREM at 75 days of age, *i.e.*, an age with no muscle denervation and no detectable motor impairment or weight loss. In mice carrying a heterozygous knock-in mutation of FUS with very slow disease progression and no progression to overt paralysis and manifest ALS symptoms, we observed the same sleep alterations at 10 months of age, while no alterations were observed at three months of age. It is striking to note that, despite the different species, and the vastly different circadian rhythms and sleep patterns, both ALS patients, ALS gene carriers and ALS mice showed increased wake and decreased NREM sleep, compared to their respective controls. The only notable species difference was that REM sleep was increased in humans and decreased in mice.

Overall, we show here a consistent pattern of ALS-associated sleep alterations, which occurs before motor impairment and before respiratory deficits. Remarkbly, sleep alterations were also observed in other neurodegenerative diseases such as Alzheimer’s (AD) (*39*) or Parkinson’s disease (PD) (*40*). However, both AD and PD patients show reductions in REM sleep which is not observed in ALS patients. Another notable difference between ALS and other neurodegenerative diseases was that, in the absence of respiratory impairment, subjective sleep quality was reported normal in ALS patients through standardized questionnaires, while in AD and PD subjective sleep is affected, suggesting that ALS patients or gene carriers are not aware of their sleep alterations. It is possible that the von Economo neurons in the anterior cingulate cortex –which are known to be affected by ALS and play an important role in subjective judgment and emotion– compromise the awareness for these alterations (*41, 42*). The sleep alterations we observe here are more similar to those observed in normal aging, and could be consistent with accelerated aging in ALS patients (*43*). Further studies on sleep microarchitecture in ALS patients are warranted to characterize similarities and differences between aging, ALS and other neurodegenerative diseases with respect to sleep alterations.

Our preclinical study also showed that alterations in key LHA signalling underlies ALS-associated sleep alterations. We previously showed that the hypothalamus is atrophied early in the disease process of ALS, even in presymptomatic gene carriers, and that ALS pathology was mostly found in the lateral hypothalamus (*10, 14, 44*). Our recent studies have shown that MCH neurons are lost in ALS mouse models and patients, while ORX neurons are preserved in animal models but lost in patients, possibly during autopsy depsite no obvious clinical correlate. MCH and ORX neurons play an important role in sleep control, with MCH neurons promoting REM sleep (*45, 46*) and ORX neurons promoting arousal (*47*). ORX neurons and MCH neurons are generally thought to antagonize each other during the different stages of sleep (*48, 49*). Since MCH neurons are lost in ALS and ORX neurons appear largely preserved, we hypothesized that the sleep alterations observed originated from an imbalance of orexinergic tone towards MCHergic output. Consistent with this hypothesis, intracerebroventricular MCH supplementation partially rescued sleep alterations in ALS mice. While sleep alterations were fully rescued in milder *Fus^ΔNLS/+^* mice, MCH rescued REM decrease, but only partially rescued wake increase, and exacerbated NREM alterations in *Sod1^G86R^* mice. Conversely, the broad ORX-antagonist entirely normalized sleep alterations in both SOD1 and FUS models. This difference of efficacy between MCH supplementation and ORX-antagonist could be due to a more prominent role of ORX neurons in sleep alterations. Yet, differences in mode of administration (i.c.v. cannulation and continuous delivery vs acute oral administration) or pharmacokinetics could also underlie such a difference. While our preclinical study demonstrates that LH neuropeptides are critical in early sleep alterations, we do not show here that these alterations originate in ORX or MCH neurons themselves, nor that other cell types or structures are not involved. Indeed, recent studies have observed involvement of other sleep controlling pathways in ALS including cholinergic pathways (*50, 51*), serotonin neurons (*52*) or more indirectly glymphatic dysfunction (*53*). Future work should perform cell specific interventions to characterize the circuit dysfunctions and their proximal causes.

The consequences of the early sleep alterations in ALS patients remain to be determined. They could be related to hypothalamic morphology and function, and therefore part of a preclinical period (*14, 44, 54*) which could not be identified in the neuroanatomical studies by Brettschneider and Braak (*55, 56*). Indeed, it could be expected that brain functions classically associated with sleep, including cognitive function and memory, could be related to sleep alterations and more generally quality of life. In addition, sleep deprivation or sleep disorders have been related to worsened motor function (*e.g.,* in PD (*40*)), postural balance (*57*) or motor skills (*36*), as well as feeding behaviour (*58*) and it would be conceivable that the sleep alterations occurring already several years prior to motor onset contributes to motor deficits or progression in ALS patients. In this setting, it would be interesting to perform a clinical trial of DORAs in ALS, not only to investigate their potential to improve sleep, but also their potential impact on cognitive deficits, weight loss or motor symptom progression that are also associated with the disease.

## MATERIALS AND METHODS

### Patients/participants

ALS patients were recruited from the inpatient and outpatient clinics of the neurologic department of the University Hospital of Ulm, Germany. The inclusion criteria for ALS patients included a diagnosis of definite ALS based on the revised El Escorial criteria (*59*). Presymptomatic carriers of fALS genes were recruited through the study centre of the Neurological University Hospital, through which first-degree relatives of confirmed familial ALS patients receive longitudinal follow-up and counselling. Controls were recruited from the general population at the neurology clinic, and matched to ALS patients based on age, sex, and geographical location; the requirement for this group was exclusion of neurodegenerative diseases. All individuals in the control group were unrelated to ALS or familial ALS.

The study in the ALS patient cohort was approved by the Ethics Committee of the University of Ulm (reference 391/18), as well as the study in the presymptomatic carriers which was also approved by the Ethics Committee of the University of Ulm (reference 68/19), in compliance with the ethical standards of the current version of the revised Helsinki Declaration. All participants gave informed consent prior to enrolment.

Medical history was documented. For ALS patients, the ALSFRS-R and characteristics of disease progression were documented (site of first paresis/atrophy, date of onset). All participants also completed validated daytime sleepiness and sleep quality questionnaires, namely the Epworth Sleepiness Scale (ESS) (*60*) and the Pittsburgh Sleep Quality Index (PSQI) (*61*).

### Mouse models

All experiments were performed in strict compliance with Directive 2010/63/EU, and new Regulation (EU) 2019/1010, and the project was reviewed and approved by the Ethics Committee of the University of Strasbourg and the French Ministry of Higher Education, Research and Innovation (Decree n°2013-118, February 1^st^, 2013). Following the “3Rs” rule, we reduced the number of included animals to a statistical minimum (as described in the statitatical section and by others (*62*)). Animals used in the experiment described in **Figure 3** were the same used describing **Figure 5**. Animal care occurred in accordance with Guide for the Care and Use of Laboratory Animals (*63*). Strict compliance with ARRIVE 2.0 guidelines has been ensured (*64*).

Wild-type FVB/NJ and FVB-Tg(Sod1G86R)M1Jwg/J (*28*) and B6-FusΔNLS1Ldup/Crl (*30, 31*) were bred in-house. From weaning age, animals were housed in same-sex sibling groups in a temperature- and light/dark-controlled environment (22°C ±2, 12:12h), starting at 0700 hours, with standard diet (A04, SAFE^®^, Augy, France) and water provided *ad libitum*. Environmental enrichment included nesting material (Nestlets, Ancare, Bellmore, NY, USA), PVC pipe, and shelter (Refuge XKA-2450-087, Ketchum Manufacturing Inc., Brockville, Ontario, Canada). Animals were kept in the animal facility until they were between 60 to 90 days old or up to 10 months for *Fus* line. Both males and females were used throughout experiments. A few days prior to the procedure, mice were separated and single-housed to habituate.

After the procedure animals were kept in the environmental control cabinet (CAB-16) at 22°C ±2, and under a light/dark cycle (12:12h) with 200 lux exposure.

### Drug

Suvorexant (FS65089) was purchased from Biosynth Ltd (Biosynth Ltd., Compton, Berkshire, United Kingdom).

### Electroencephalography in patients and subjects

All participants, ALS patients, healthy controls, fALS gene carriers and fALS controls underwent a 1-night full polysomnography, involving monitoring of various physiological parameters including electroencephalogram (EEG), surface electromyogram (EMG), electrooculogram (EOG), respiratory effort and flow, pulse and oxygen saturation. All measurements were conducted according to the criteria of the American Academy of Sleep Medicine (AASM) guidelines (*65, 66*). The EEG electrodes were placed according to the international 10-20 system, the following electrodes were used in each subject: Fz, C3, C4, Cz, P3, P4, Pz, O1, O2, A1, and A2. The sampling rate was 512 Hz in each case. The individually different point in time at which the participant turned off the lights and tried to sleep was marked with a “lights off” marker in each recording.

### Sleep analyses in patients and subjects

Analyses were performed using Python scripts (only compatible with Python 3.10 or newer, Python Software Foundation. Python Language Reference, version 3.12. Available at http://www.python.org) relying on MNE package (*67*). EEGs were first de-identified using the open-source Prerau Lab EDF De-identification Tool (Version 1.0; 2023) in Python (Prerau Lab EDF De-identification Tool [Computer software], 2023, Retrieved from https://sleepeeg.org/edf-de-identification-tool). De-identified EEGs were then notch-filtered to remove the 50Hz powerline. Independent component analysis was performed to remove all remaining artefacts from the signal (*68–72*). Analyses were limited to both sensorimotor cortices (C_3_ and C_4_), which are known to be impaired at the onset of the disease, as well as nearby interhemispheric sulci (Fz, Cz and Pz). Sleep staging was performed on a 6-hour window with 30 seconds epochs, starting when lights were turned off, using YASA deep learning algorithm (*26*). The automated sleep staging, hypnograms, and spectrograms were performed using Welch’s method (*73*). Sleep pressure was determined using the area under the curve (AUC) of Delta power (0.5-4Hz) of the first hour of the 6-hour window. Simpson’s rule was used to compute the AUC. REM efficiency was computed by dividing Theta power (4-8Hz) by Delta power (0.5-4Hz) specifically during REM epochs. Sleep staging and analysis were performed following the AASM’s guidelines (*65, 66*).

### Wireless electrocorticography and analyses in mouse models

Animals were maintained under deep anaesthesia via inhalation of isoflurane (Baxter). Anaesthesia was monitored by periodic paw withdrawal and respiratory reflexes throughout the procedure. Amid the procedure, a single injection of Meloxicam injectable (Metacam, 10 mg/mL; Boehringer Ingelheim) and atropine sulfate (Atropine, 0.5 mg/mL, Aguettant) was given subcutaneously to the animal. The fur was trimmed, and the skin cleaned thoroughly using sterile swabs of 2% chlorhexidine and 70% alcohol solution. A single local subcutaneously injection of Anhydrous hydrochloride (Bupivacaine, 2 mg/kg, Mylan) and Lidocaine (Lurocaine, 2 mg/kg, Vetoquinol) under the skull’s skin was applied. Anaesthetised animals were placed in a stereotaxic frame (World Precision Instruments) and draped in sterile sheets. Three small holes were drilled using a micro-drill (Cordless Microdrill, RWD, San Diego, CA, USA) in the skull to permit the insertion of two leads at those stereotaxic coordinates: Negative electrode: Bregma +1.00mm, midline +1.00mm; Positive electrode: Bregma -3.00mm, midline -3.00mm.

Two additional leads were implanted intramuscularly in the neck’s muscle. The sterile wireless implant (HD-X02 dual biopotential transmitter, Data Science International Inc., St. Paul, MN, USA) was implanted subcutaneously above the scapulae, as well as the osmotic pump for the MCH experiments. The skin was stitched using a sterile suture (Ethicon 5-0 Vicryl^™^ Plus coated with antibacterial substance) and N-butyl-cyanoacrylate (Surgibond) to reinforce the stitching in some areas. Finally, animals received a single subcutaneous injection of injectable Buprenorphine (Buprecare, 0.3 mg/mL, Axience) to prevent any possible pain later, as well as a shot of sterile and warm Sodium chloride for infusion (0.9%). The animals were constantly on a heating pad throughout the whole surgery to monitor their body temperature using a rectal probe. Post-surgery, Meloxicam (Metacam, 5 mg/kg, Boehringer Ingelheim) was given in the drinking water and GelDiet^®^ Recovery (ClearH2O) as a wet source of nutrients for a better recovery of the animals.

For MCH experiments prior to the electrocorticography procedure, Azlet^®^ Osmotic Pumps (Azlet, Cupertino, USA) were loaded with either the vehicle (sterile sodium chloride 0.9%) or MCH (human MCH, #3806, Tocris Biosciences) 0.1mg/mL diluted in the vehicle solution) and attached to the Brain Kit Infusion 3 with 2 adjustment spacers (Azlet, Cupertino, USA). The brain cannula was implanted into the right lateral ventricle at the following coordinates: Bregma -0.8mm, midline +0.4mm, as previously described (*14*). Pumps delivered, either the vehicle or MCH solution, at a constant rate of approximately 1.2 µg/day for 15 days.

For Suvorexant experiments, drug administration was done by gavage, DMSO 100% (Fluka, 0.5mL/mL) as vehicule, and finally with Suvorexant (20mg/kg diluted in the vehicle solution).

In both cases, after the recovery period, the animals were recorded using Ponemah 6.51 acquisition software (Data Science International Inc., St. Paul, MN, USA) for 24 hours for baseline. The animals were able to move freely in their home cage using this novel implantable and wireless telemetry device. Recording sessions began between 0830–0900.

### Electrocorticography analysis in mice

We used NeuroScore software for sleep and seizure analysis 3.4 (Data Science International Inc., St. Paul, MN, USA) to analyse and score the EEG, EMG and activity count of the animals at baseline, with the vehicle and with Suvorexant, or MCH. For both vehicle and Suvorexant or MCH, the hour following the gavage was removed from the recording to ensure minimal stress effect would be seen on the recording due to the gavage.

EEG and EMG signals, as well as activity counts, were used to score sleep/wake behavioural states using NeuroScore software for sleep and seizure analysis 3.4 (Data Science International Inc., St. Paul, MN, USA) Rodent Sleep automated scoring method. We used 5sec epochs for both the EEG and EMG. Then the automated scoring method classified each epoch into one of the following behavioural states: (0) wake (low-voltage fast EEG, high-voltage EMG, with frequent activity counts); (1) non-rapid eye movement 1 (NREM1 sleep: spindling and high-voltage EEG with slow waves, low-voltage EMG and no activity counts); (2) non-rapid eye movement 2 (NREM2 sleep: spindling and high-voltage EEG with slow waves, very low voltage EMG) and no activity counts; (4) rapid eye movement (REM sleep: low-voltage and fast EEG combined with very low-voltage EMG, with occasional short-duration, large-amplitude EMG activity due to muscle twitches as well as sporadic short-duration activity counts).

The activity count was used as a double-check for false-positive EMG activity. Wake, REM and NREM were analysed as a percentage over the 24-hour recording period, with epochs of 5 seconds.

Data were extracted from NeuroScore and used in combination with a Python script (Python Software Foundation. Python Language Reference, version 3.12. Available at http://www.python.org) to further process the data, and an already available Python toolbox, YASA (*26*), to generate hypnograms. Hypnograms were generated following the AASM’s guidelines (*65, 66*).

### Immunofluorescence in mouse models

Mice were euthanized with a lethal i.p. injection of 120mg/kg of pentobarbital sodium and transcardially perfused using 0.01M phosphate buffer saline (PBS) followed by 4% paraformaldehyde (PFA) in 0.01M phosphate buffer. Brains were post-fixed overnight in 4% PFA at 4°C. Brains were then bathed in PBS with 30% Glucose for 72 hours, after which those were bathed in isopentane at -40°C and stored at -80°C. Brain slices (thickness 30μm) were done using a CM3050S cryostat (Leica Biosystem, Wetzlar, Germany) and collected in PBS with 0.02% Azide. Sections were selected using the Allen Mouse Brain Atlas (Allen Reference Atlas Mouse Brain [brain atlas]. Available from atlas.brain-map.org). A panel of 5 sections from the region of interest were stained against ORX and were anatomically matched with their respective controls. Sections were first blocked in PBS with 5% horse serum and 0.5% Tritton-X-100 for 1 hour at room temperature (RT). The primary antibody was incubated for 2 hours at RT ORX (SC-80263, 1:250, Santa Cruz Biotechnology, Santa Cruz, CA, USA) diluted in PBS. Sections were washed thrice in PBS and incubated for 1 hour at RT with a fluorescent conjugated secondary antibody (711-605-152, 1:1000, Jackson ImmunoResearch), diluted in PBS. Finally, sections were washed thrice in PBS before being mounted on microscope slides using VectaShield Plus with DAPI (H-200010, Vector Laboratories). Imaging was performed using an Axio Imager M2 epifluorescence microscope (Zeiss Laboratories, Thornwood, NY, USA). Tiles were merged into a 2D plan for further quantification using Zen Pro software (Zeiss Laboratories, Thornwood, NY, USA). Image quantification was performed using Fiji (*74*). All quantifications were achieved by an unbiased investigator blinded to genotype or disease status.

### Patients’ inclusion process

The exclusion criteria were intended to exclude all possible circumstances that might otherwise alter sleep architecture. For this reason, participants who had an apnoea-hypopnea index (AHI) above 20 per hour or participants who had a periodic limb movement index (PLMSI) above 50 per hour were excluded. In particular, we intended to exclude respiratory insufficiency in ALS patients. Respiratory insufficiency develops earlier or later in the progression of ALS, depending on the individual course, but is generally present in advanced stages, and is known to influence sleep architecture. For this reason, ALS patients received transcutaneous capnometry in addition to polysomnography.

**Table 3.**
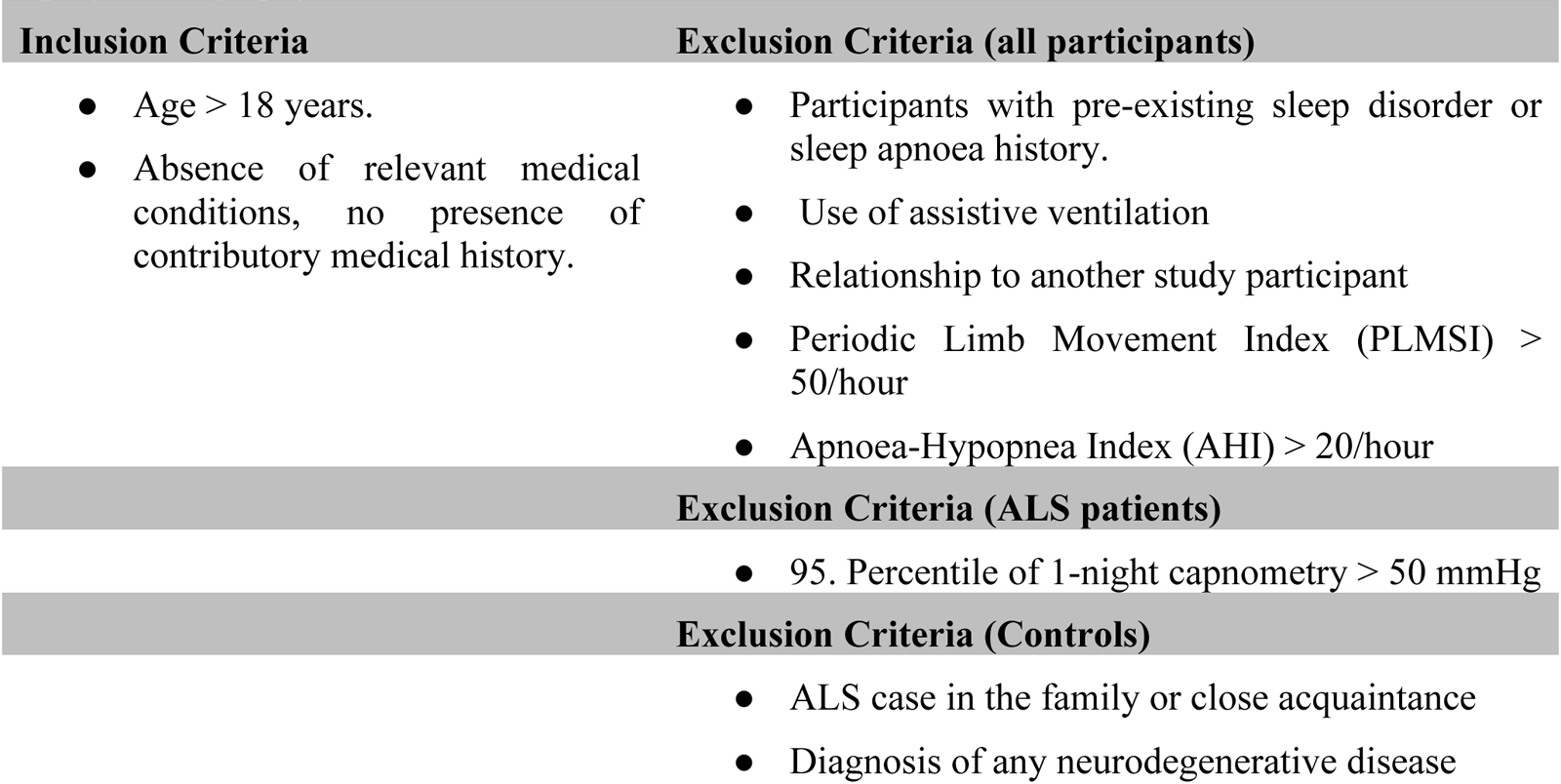
Inclusion and exclusion criteria for the cohort of ALS patients and healthy controls.

**Table 4.** Inclusion and exclusion criteria for the cohort of first-degree relatives of fALS cases.

| Inclusion Criteria | Exclusion Criteria |
| --- | --- |
| Age > 18 years.<br>Absence of relevant medical conditions, no presence of contributory medical history.<br>First-degree relation to ALS Patient with confirmed mutation leading to familial ALS. | Participants with pre-existing sleep disorder or sleep apnoea history.<br>Use of assistive ventilation<br>Relationship to another study participant<br>Periodic Limb Movement Index (PLMSI) > 50/hour<br>Apnoea-Hypopnea Index (AHI) > 20/hour<br>Diagnosis of any neurodegenerative disease |

### Statistical analyses

G*Power software (Version 3.1.9.6 for macOS; 2023) was used to determine sufficient sample size needed to reach significant statistical power using an *A priori* Student’s t-test coupled with a linear bivariate regression (*75, 76*).

Prior to any statistical analysis, normality and homoscedasticity were both tested respectively with Shapiro-Wilk test (*77*) and Bartlett’s test (*78*).

Statistical analysis of two groups were performed using an independent Student’s t-test, from Pingouin (*79*), using the Welch t-test correction, from SciPy, as recommended by Zimmerman DW 2004 (*80*) and with a large Cauchy scale factor due to the considerate effect size (*81*).

For statistical analysis of three or four groups, a One-way ANOVA or Two-way ANOVA was performed using Pingouin (*79*) toolbox. For both One-way ANOVA and Two-way ANOVA, a one-step Bonferroni correction (*82*) was applied. We evaluated whether sex-specific effect was present in all our analysis by performing a Two-way ANOVA with a one-step Bonferroni correction (*82*) for both sex. Sex was self-reported in both ALS cohorts.

Simpson’s rule (*83, 84*), from SciPy (*85*), was used to determine the AUC. PCAs were performed using scikit-learn package (*86*).

Data are presented as violin plots with all points and expressed as average ± interquartile. Plots were generated using Seaborn and Matplotlib packages (*87, 88*). Results were deemed significant when their adj. p_value_<0.05. Here, only corrected p_value_ (adj. p_value_) are shown.

## Code availability

Scripts used to perform the analysis are available upon reasonable request at: https://github.com/sjg2203

## Data Availability

All data produced in the present study are available upon reasonable request to the authors

## Acknowledgements

The authors thank the patients and their families, as well as volunteers for the participation in these studies. The authors are thankful to the Animal Facility Core (PEFRE, University of Strasbourg, INSERM, UMS 38) and to Claudia de Tapia for technical assistance. The Microscopy Core (PIC-STRA, University of Strasbourg, INSERM, UMS 38), and in particular Pascal Kessler, for the imaging assistance. Daniel Beckett for proof reading the final manuscript.

## Author contributions

Conceptualization: SJG, CL, MS, GSL, ACL, LD, MB

Methodology: SJG, CL, PHL, ACL, MB

Investigation: SJG, CL, AK, PW, JD, JK, KK, SA, MB

Formal analysis: SJG, CL, ACL, LD, MB

Visualization: SJG, CL

Resources: SJG, CR, PHL, ACL, LD, MB

Funding acquisition: ACL, FR, LD

Project administration: AK, PW, JD, FR, ACL, LD, MB

Supervision: ACL, LD, MB

Writing – original draft: SJG, CL, LD, MB

Writing – review & editing: all authors.

