## Supplementary material for "Lateral hypothalamus drives early-onset sleep alterations in amyotrophic lateral sclerosis": 5 Supp Figures and 2 Supp Tables

**List of Supplementary Materials.**

Figure S1 to S5

Table S1 to S2

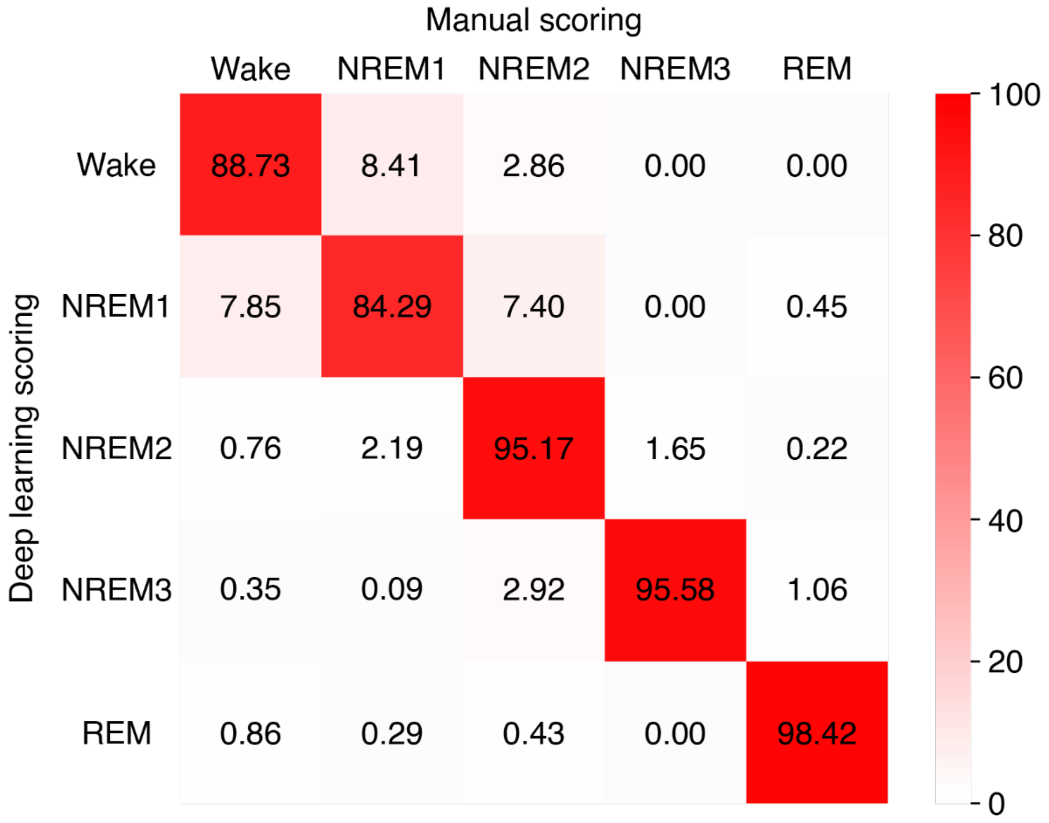


**Figure S1. Sleep stage concordance between deep learning and manual scoring in early ALS patients.**
Values are shown as percentage of the row, n=8 recordings; 4 controls (2 women, 2 men), 4 ALS patients (2 women, 2 men).

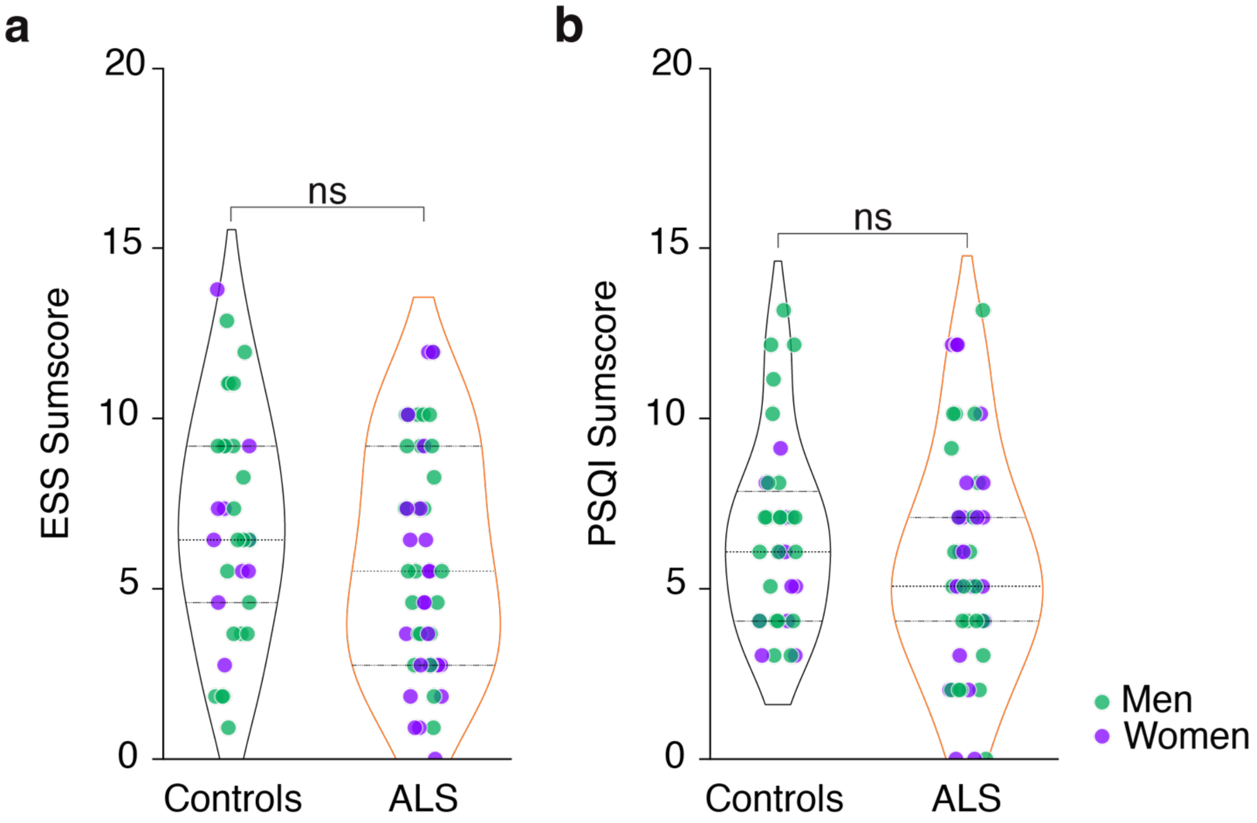


**Figure S2. Sleep questionnaires in early ALS patients.**

(**a**) Epworth Sleepiness Scale (ESS) Sumscore of healthy controls and ALS patients.

(**b**) Pittsburgh Sleep Quality Index (PSQI) Sumscore of healthy controls and ALS patients.

In all panels, men are shown in green and women in purple.

Not significant (ns) adj. $p_{value}>0.1$, independent Student's t-test with Welch's t-test correction; sex effect adj. $p_{value}=0.2599$. Data are presented as median and interquartile ranges. Corrected $p_{value}$ are shown.

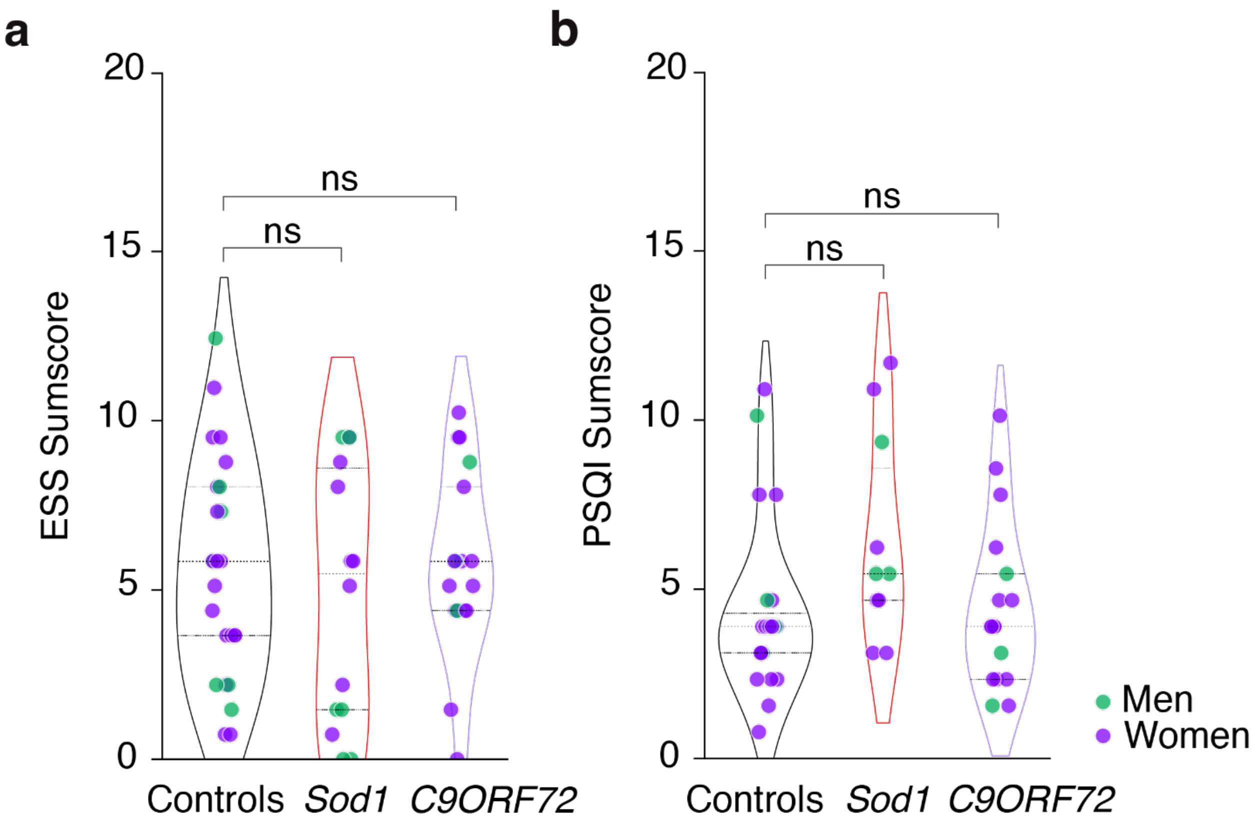


**Figure S3. Sleep questionnaires in presymptomatic ALS gene carriers.**

(**a**) Epworth Sleepiness Scale (ESS) Sumscore of fALS controls and presymptomatic gene carriers.

(**b**) Pittsburgh Sleep Quality Index (PSQI) Sumscore of fALS controls and presymptomatic gene carriers.

In all panels, men are shown in green and women in purple.

ns adj. $p_{value}$>0.1, One-way ANOVA with one-step Bonferroni correction; sex effect adj. $p_{value}$=0.5131. Data are presented as median and interquartile ranges. Corrected $p_{value}$ are shown.

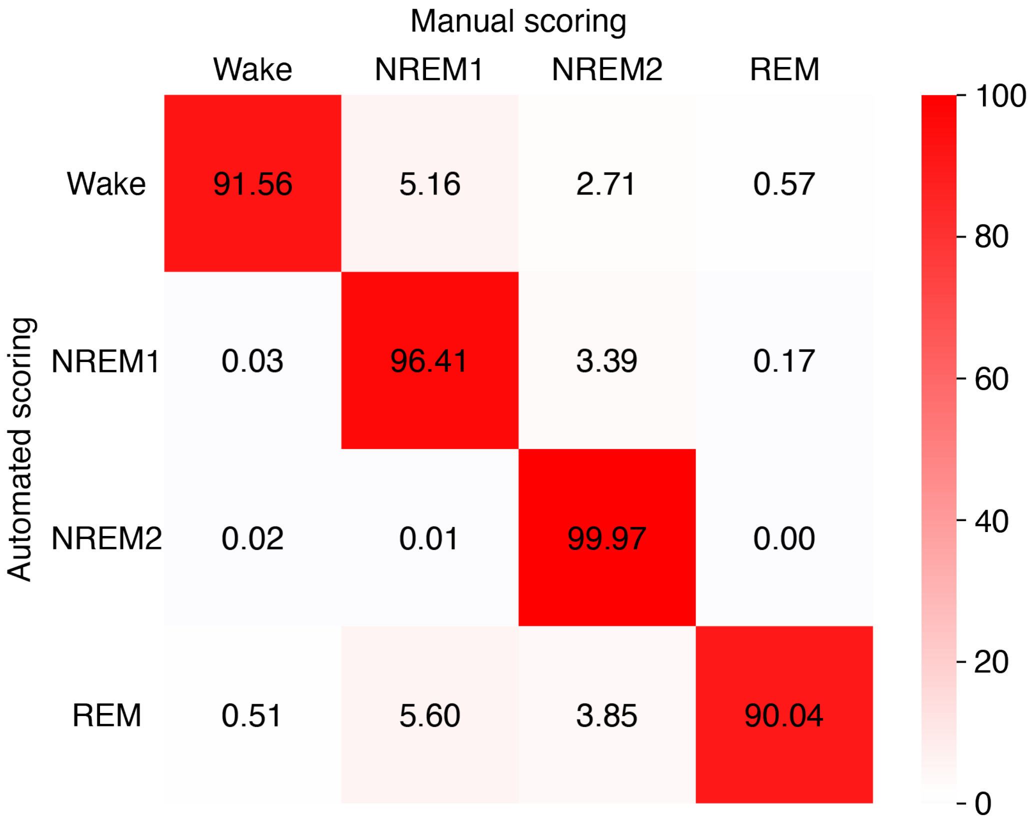


**Figure S4. Sleep stage concordance between automated and manual scoring in mouse models of ALS.**

Values are shown as percentage of the row, n=6 recordings; 3 female WT mice, 3 female *Sod1*$^{G86R}$ mice.

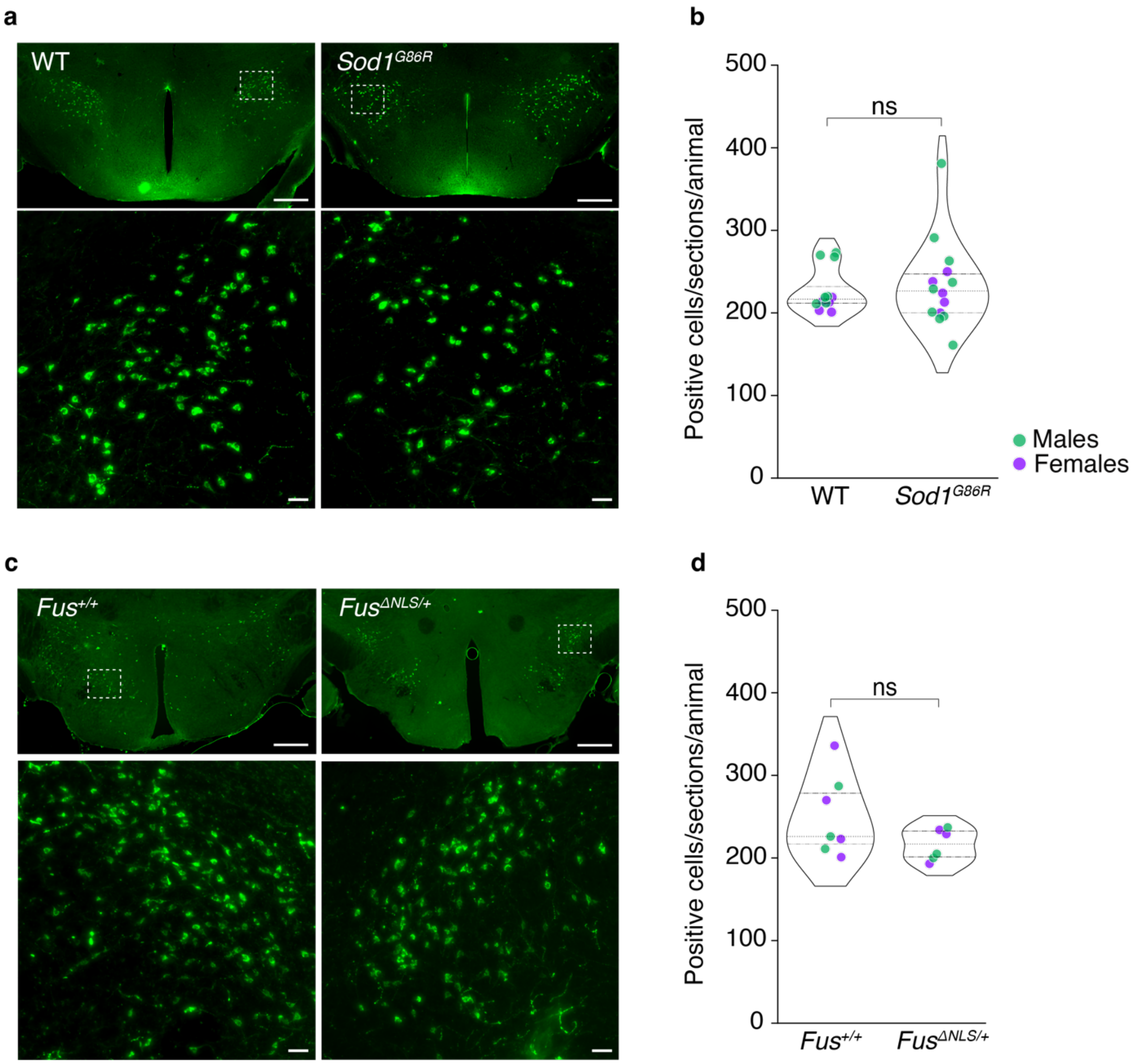


**Figure S5. Orexin counts remains unchanged in presymptomatic ALS mouse models.**

(**a**) Representative ORX immunostaining in *Sod1*$^{G86R}$ or WT littermates (75 days of age).

(**b**) Number of ORX-positive cells per sections in *Sod1*$^{G86R}$ or WT littermates. *Sod1*$^{G86R}$ mice: n=12; 5 females and 7 males; WT: n=14; 5 females and 9 males.

(**c**) Representative ORX immunostaining in *Fus*$^{\Delta NLS/+}$ or *Fus*$^{+/+}$ littermates (10 months of age).

(**d**) Number of ORX-positive cells per sections in *Fus*$^{\Delta NLS}$ or *Fus*$^{+/+}$ littermates. *Fus*$^{\Delta NLS/+}$ mice: n=7; 4 females and 3 males; *Fus*$^{+/+}$: n=6; 3 females and 3 males.

Lower panels display a greater magnification of the region of interest indicated by the dashed rectangle, with a scale is 50μm. The upper panels have a 500μm scale.

ns adj. $p_{value}$>0.05, independent Student's t-test with Welch's t-test correction; sex effect *Sod1*$^{G86R}$ adj. $p_{value}$=0.4072, *Fus*$^{\Delta NLS/+}$ adj. $p_{value}$=0.3649. Data are presented as median and interquartile ranges. Corrected $p_{value}$ are shown.

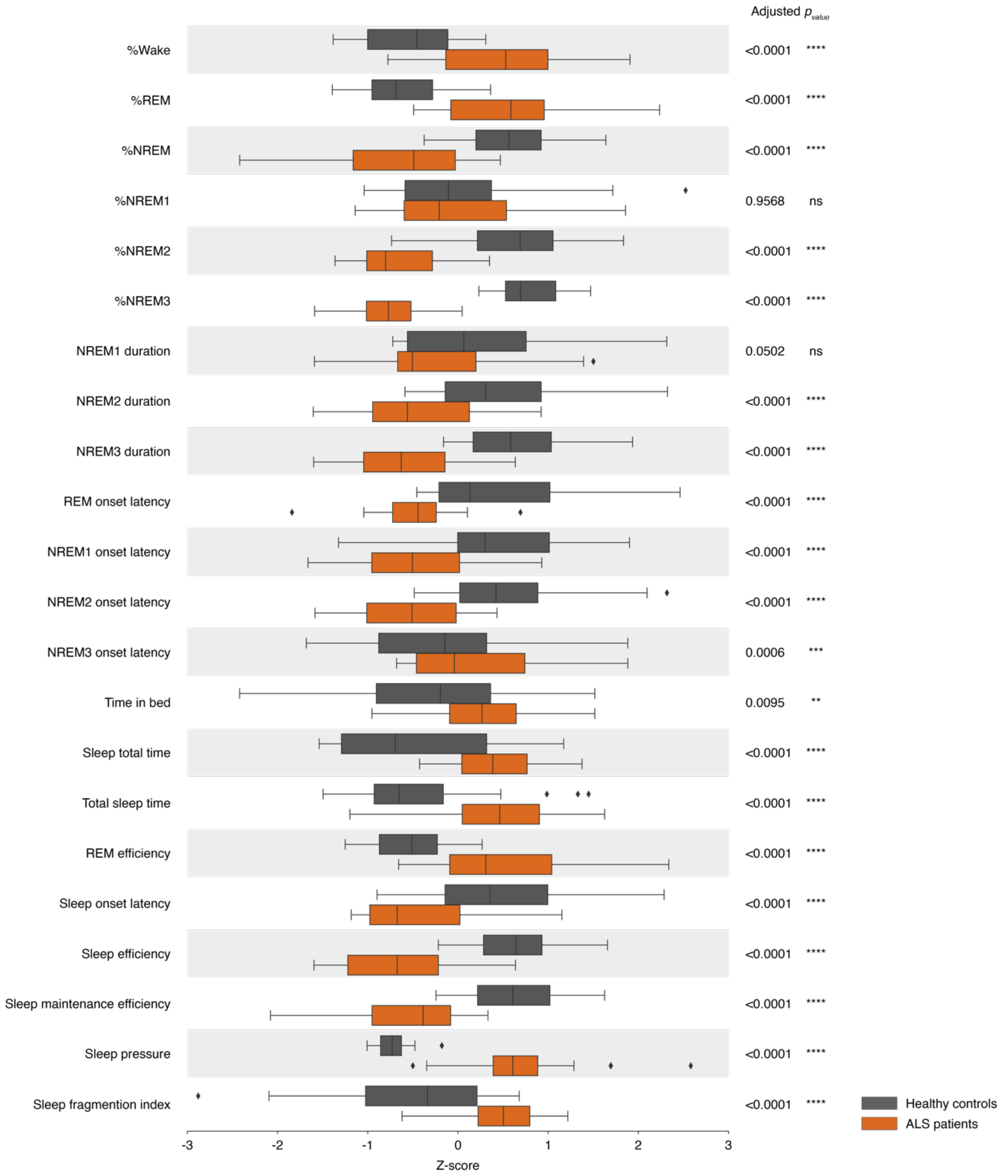


**Table S1. Complementary sleep alterations in early ALS patients.**

** adj. $p_{value}$<0.01, independent Student's t-test with Welch's t-test correction. Data are presented as mean and 95% confidence intervals. Corrected $p_{values}$ are shown.

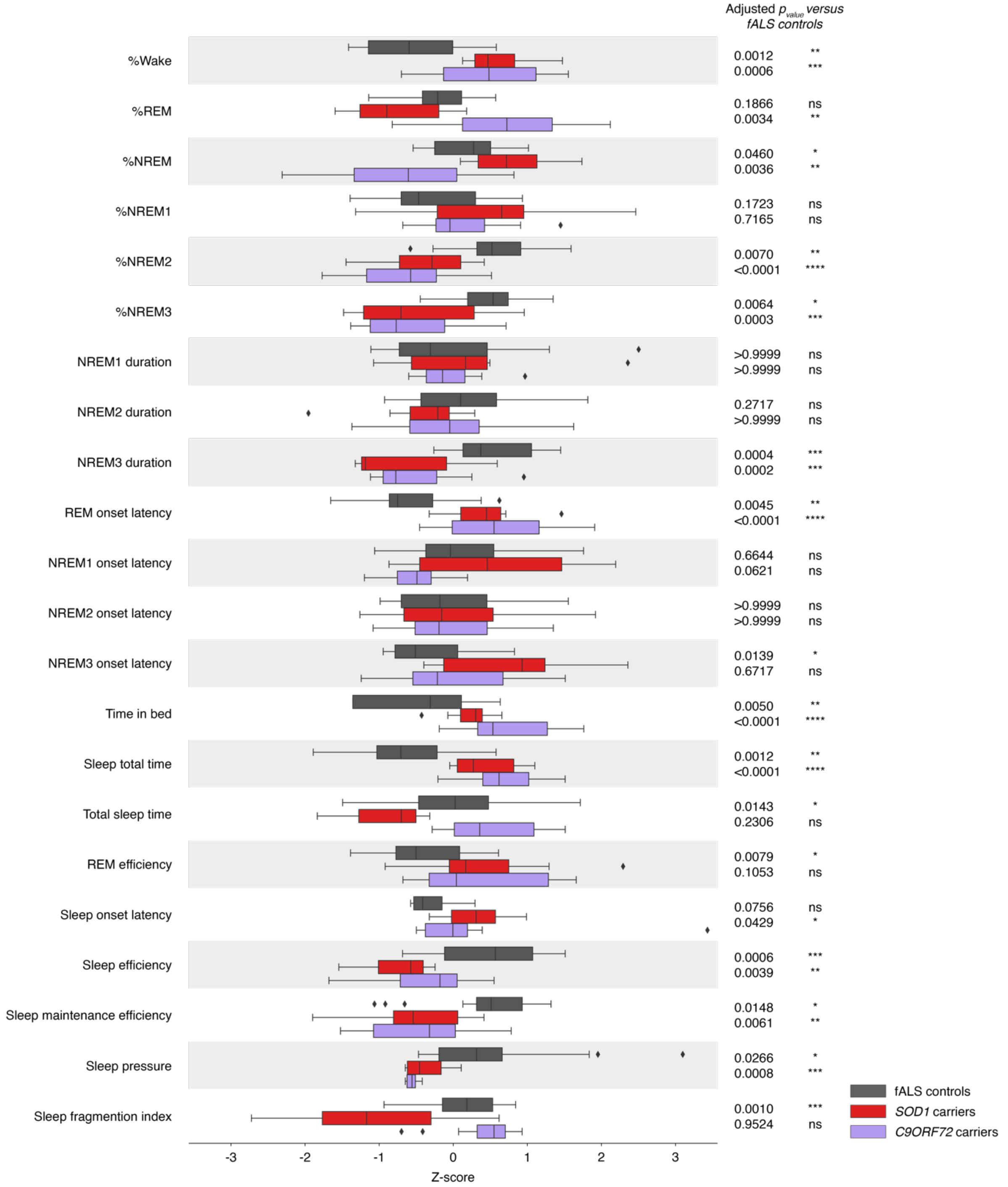


**Table S2. Complementary sleep alterations in presymptomatic ALS gene carriers.** * adj. $p_{value}<0.05$, One-way ANOVA with one-step Bonferroni correction. Data are presented as mean and 95% confidence intervals. First adj. $p_{value}$ represents healthy individuals versus *SOD1* carriers, the second adj. $p_{value}$ represents healthy individuals versus *C9ORF72* carriers. Corrected $p_{values}$ are shown.
